# Additional tuberculosis case detection through repeat household contact investigation at 6 and 12 months: a retrospective cohort study in Kasama, a peri-urban district in Zambia

**DOI:** 10.64898/2026.11.05.26367344

**Authors:** Phallon Blessing Mwaba, Mutale Mukupa, Josaya Sakala, Innocent Mwaba, Daniel Siameka, Minyoi Mubita Maimbolwa, Chitalu Chanda, Muhammad Mputu, Kevin Mabvuto Zimba, Nancy Kasese-Chanda, Rhehab Chimzizi, Angel Mubanga, Monde Muyoyeta, Mary Kagujje

**Author notes:** Corresponding author: **E-mail:** (PBM).

## Abstract

**Background:** Tuberculosis contact investigation (TBCI) facilitates early detection of TB among household contacts. However, evidence on the additional contribution of repeat TBCI in high-burden settings remains limited. We assessed the contribution of repeat TBCI at 6 and 12 months to TB case detection in Kasama District, Zambia.

**Methods:** We conducted a retrospective observational cohort analysis of routinely collected programme data from five purposively selected high-volume TB diagnostic facilities. Household contacts of bacteriologically confirmed TB patients notified between January 2023 and April 2024 underwent symptom-based TB screening at baseline and at 6 and 12 months. We estimated round-specific TB detection yields, the cumulative proportion diagnosed with TB, and the contribution of repeat investigations to overall case detection.

**Results:** Among 1,250 index TB patients, 3,648 household contacts were identified and screened at baseline. TB was diagnosed in 143 (3.9%) contacts at baseline, 31 (0.9%) at month 6, and 25 (0.7%) at month 12. Overall, 199 (5.5%) contacts were diagnosed with TB across the three screening rounds, of whom 159 (79.9%) had bacteriologically confirmed disease. Repeat investigations identified 56 additional cases, representing 28.1% of all TB diagnoses.

**Conclusions:** Repeat TBCI at 6 and 12 months identified additional TB cases beyond baseline screening, although detection yields declined across successive rounds. These findings support further evaluation of risk-based follow-up strategies and more sensitive baseline screening approaches to optimize TB case detection and resource utilization in high-burden settings.

## Introduction

Tuberculosis (TB) remains a significant public health challenge in Zambia; an estimated 58,000 people developed TB, and approximately 4100 died from the disease in 2024 (1). In the same year, approximately 19% of Zambia’s estimated TB cases were not captured through routine TB notification systems, representing about 11,000 people who developed TB but were not diagnosed and/or officially notified (1). To reduce the TB burden, TB patients should be identified, treated early and their close contacts traced and evaluated for TB (2).

TB contact investigation (TBCI), a form of active TB case finding (3,4), has the potential to facilitate early identification of undiagnosed TB cases and timely initiation of treatment (5,6). Early treatment has the potential to reduce community TB spread. Studies have consistently demonstrated that effective TBCI can significantly reduce community TB transmission and improve outcomes in both high- and low-burden countries, hence key to achieving global TB elimination targets (7). Additionally, TBCI serves as an entry point for preventive measures such as TB preventive treatment (TPT) (5) to help accelerate reduction in TB incidence (8).

Traditionally, TBCI has emphasized a single round of contact investigation, particularly prioritizing children younger than five years who are contacts to bacteriologically confirmed TB patients. Recent evidence increasingly supports the potential value of repeat TBCI at defined intervals (9). One of the few African studies evaluating repeat contact investigation in Africa, found a 2.3% cumulative yield among TB contacts evaluated (10). While there is a paucity of local and regional evidence, a prospective study in the United States and Canada involving 4,490 close contacts of 718 patients with culture-confirmed pulmonary TB established that 44% of the total yield from TBCI came from follow ups at 3, 6, and 12 months following notification of the index patient (11). This aligns with evidence that TB infection can remain latent for prolonged periods, with the risk of progression being disproportionately concentrated in the first two years after infection (3,4). This risk is particularly relevant among household contacts of people with TB, who have substantially higher risks of TB infection and disease than the general population (12).

Despite this evidence, there is limited evidence on the feasibility and programmatic value of systematic repeat TBCI in high-TB-burden settings such as Zambia. Zambia incorporated repeat contact investigation into its 2022 Consolidated TB Guidelines, recommending repeat investigation at six months, corresponding to the completion of treatment for the index patient, and subsequently at 12 and 24 months. Despite this provision, implementation in routine practice has been slow. We analysed routinely collected programme data from the pilot implementation of repeat TBCI in one district in northern Zambia to assess its contribution to additional TB case identification..

## Methodology

### Study Design

We conducted a retrospective observational cohort analysis of routinely collected de-identified programmatic data from household contacts of bacteriologically confirmed TB patients. The data were captured in Ministry of Health registers during a programmatic implementation of repeat TBCI between January 2023 and April 2024.

### Study sites and study population

In 2023, the USAID funded Tuberculosis Local Organizations Network (TBLON) project commenced a phased implementation of repeat TBCI in Northern Province-Kasama district. According to the 2022 census estimates, Kasama District Health office estimated the district catchment population of about 348, 552 in 2023 (13). In 2023, the district notified a total of 1,454 TB cases from all health facilities (417 per 100 000 population) (unpublished programme data).

Of the ten diagnostic sites in the district, we included a total of five high volume TB diagnostic facilities: four primary healthcare facilities and one second level (general) hospital. Facilities were purposively selected as they were the only sites where repeat TBCI had been introduced during the early implementation phase. The study population comprised all household contacts (children and adults) of bacteriologically confirmed TB patients notified between January 2023 and April 2024. The April 2024 cutoff allowed all enrolled contacts to reach the 12-month follow-up point before data extraction in May–July 2025. As the study aimed to enroll all eligible contacts within this defined period, a formal sample size calculation was not conducted.

### Procedures

During the implementation period of repeat TBCI, a structured approach was followed. Once notified, TB patients were paired with community health workers (CHWs) (Peer-Patient Pairing) to provide health education, adherence support and conduct home visits for repeat TBCI. Home visits were conducted to evaluate contacts for TB at months 0 (initial contact investigation around the date of TB diagnosis for the index patient), 6 (corresponding approximately to completion of treatment of the index patient) and 12 (additional follow-up recommended by the national guidance) after the notification of the index case. To ensure comprehensive contact follow-up, individuals who had relocated were either traced physically or screened remotely on phone and symptomatic ones referred to the nearest health facility for further screening and testing. For all the options above, the WHO four TB symptoms screening (cough, fever, weight loss and night sweats) was conducted on all contacts at each time point and only those symptomatic for TB were requested to submit samples for laboratory evaluation using GeneXpert. Due to the limited access to chest X-ray, contact screening did not include mandatory chest x-rays.

At the close of each work week, chest clinic In-charges reviewed the contact tracing registers to identify index TB patients due for repeat TBCI in the subsequent week. The patients identified were assigned to the same CBVs who had previously been paired with the corresponding index TB patient for contact investigation during that week. If the paired CBV was unavailable, chest clinic in charges assigned alternative CBVs. The CBVs submitted TBCI reports to the chest clinic in-charges and the Community Mobilization Officer (CMO) at the end of each week. Registers were updated by CBVs upon completion of their tasks. If established that contacts had relocated from their documented address, an initial remote symptom screening was conducted on phone and if the patient was symptomatic, they were linked to the nearest health facility for detailed clinical review and investigations. Whenever possible, HCWs contacted the nearest health facility to the place where contacts had relocated to facilitate follow-up, and findings of the screening shared with them.

We collected data from treatment and contact tracing registers between May and July 2025 using a pretested electronic tool designed in DHIS2 tracker. Health Care Workers in the Ministry of Health facilities extracted de-identified individual level data on contacts to bacteriologically confirmed TB index patients and the index TB patients themselves. A Data Coordinator and a Clinical Mentor verified the data by randomly sampling 10% of the index TB patients’ contacts comparing data collected with that in the registers to ensure quality data.

### Data analysis

The data collected using a DHIS2 tracker-based electronic tool was extracted into Excel. After data cleaning, data was exported to Stata version 18.5 for analysis. We used descriptive statistics to summarise the characteristics of TB contacts by reporting frequencies and percentages for categorical variables. The contact tracing ratio was determined by calculating the median number of contacts per index case with the interquartile range (IQR). TB yield at different follow-on time points (months 0, 6 and 12) was calculated as the proportion of contacts diagnosed with TB relative to those screened/investigated. The contribution of follow-on TBCI was assessed by determining the proportion of TB cases identified at each time point relative to the total TB cases detected over the study period.

### Ethical Considerations

Ethics approval for the study was obtained from University of Zambia Biomedical Research and Ethics Committee, Lusaka, Zambia (Reference number 1773–2021) to analyse existing programme data. The University of Zambia Biomedical Research and Ethics Committee, Lusaka, Zambia (Reference number 1773–2021) also waived the requirement of written informed consent, as the analysis was retrospectively performed using routine data which was de-identified before analysis.

## Results

A total of 1,250 bacteriologically confirmed index TB patients were enrolled, from whom 3,648 household contacts were elicited and all the contacts were screened at baseline (Month 0) (Fig 1) and followed up for 1 year. Among contacts screened at baseline, 3,505 were TB-negative and eligible for repeat TBCI at Month 6, while 3,474 remained eligible and were investigated at Month 12. At baseline, 143 contacts (3.9%) were diagnosed with TB. Of these, 106 (74.1%) were bacteriologically confirmed and 37 (25.9%) were clinically diagnosed. At Month 6, 31 new TB cases (0.9%) were identified, including 28 (90.3%) bacteriologically confirmed and 3 (9.7%) clinically diagnosed cases. At Month 12, 25 additional TB cases (0.7%) were detected, all (100%) bacteriologically confirmed. In total, 199 TB cases (5.5%) were detected among all contacts across the three screening rounds, of which 159 (79.9%) were bacteriologically confirmed.

**Figure 1:**
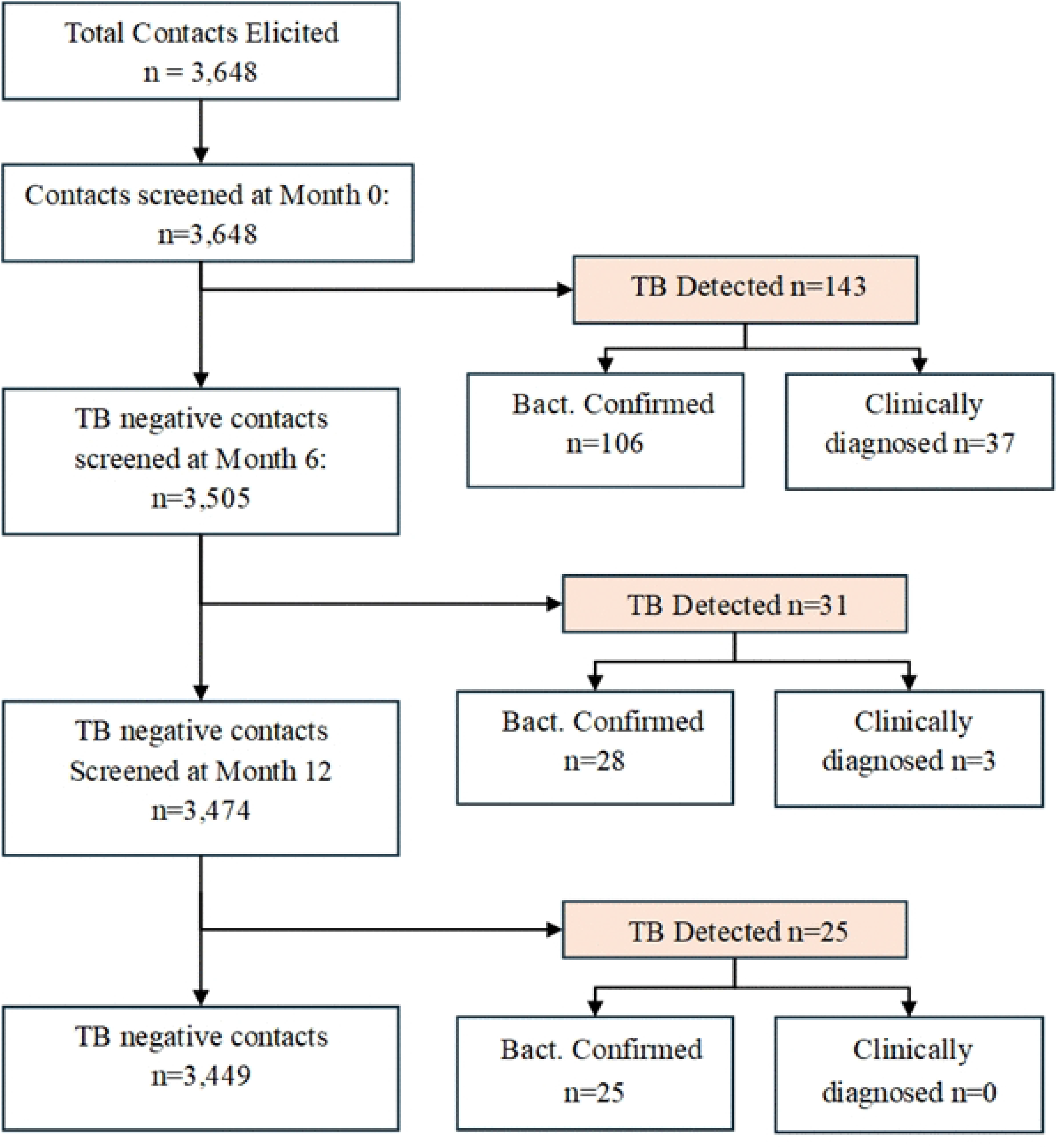
Participants flow diagram for repeat contact investigation

### Characteristics of contacts

Of the 3,648 contacts screened for TB at Month 0, 48.9% (n=1,783) were male, the median age was 35 years; IQR: 21–50 years, 83.9% (n=3,060) were aged ≥15 years, and 0.7% (n=27) were HIV positive. The median contact density (number of contacts per household) analysis was 3; IQR:1-4 with 33.5% (n=1,222/3,648) of the contacts screened came from households with ≥5 contacts, while 21.1% (n=769/3,648) were from households with ≤2 contacts.

While females accounted for 53.9% (n=77/143) of the TB cases detected at month 0, males constituted 58.1% (n=18/31) at month 6 and (52.0%, n=13/25) at month 12. Children aged under 15 years accounted for 16.1% (n=32/199) of all TB cases detected, comprising 16.8% (n=24/143), 6.5% (n=2/31), and 24.0% (n=6/25) of cases identified at months 0, 6, and 12, respectively. Among the total TB detected across months 0, 6 and 12, 95.5% (n=190/199) were HIV negative (93.7%, 100% and 100% at months 0,6 and 12 respectively). The cumulative proportion of contacts diagnosed with TB was highest among those from households with two or fewer contacts (10.7%, n=82/769). Whereas contribution to TB notifications from households with ≤ 2 contact declined over time (48,3%, 35.5% and 8% at months 0, 6 and 12 respectively), contributions from households with ≥3 contacts increased progressively (51.7%, 64.5% and 92% at months 0, 6 and 12 respectively).

**Table 1:** Characteristics of elicited contacts and TB detected at baseline and repeat time points (n=3,648)

| **Characteristics** | **Total Contacts (n=3,648)** | **Month 0** |  | **Month 6** |  | **Month 12** |  |
|---|---|---|---|---|---|---|---|
|  |  | TB (n=143) | No TB (n=3,505) | TB (n=31) | No TB (n=3,474) | TB (n=25) | No TB (n=3,449) |
| Sex, *n (%)* |  |  |  |  |  |  |  |
| Male | 1,783 (48.9) | 66 (46.2) | 1,717 (49.0) | 18 (58.1) | 1,699 (48.9) | 13 (52.0) | 1,686 (48.9) |
| Female | 1,865 (51.1) | 77 (53.9) | 1,788 (51.0) | 13 (41.9) | 1,775 (51.1) | 12 (48.0) | 1,763 (51.1) |
| Age in years (median [IQR]) | 35 [21-50] |  |  |  |  |  |  |
| <15; *n(%)* | 588 (16.1) | 24 (16.8) | 564 (16.1) | 2 (6.5) | 562 (16.2) | 6 (24.0) | 556 (16.1) |
| ≥15; *n(%)* | 3,060 (83.9) | 119 (83.2) | 2,941 (83.9) | 29 (93.5) | 2,912 (83.8) | 19 (76.0) | 2,893 (83.9) |
| HIV status *n(%)* |  |  |  |  |  |  |  |
| Positive | 27 (0.7) | 9 (6.3) | 18 (0.5) | 0 (0.0) | 18 (0.5) | 0 (0.0) | 18 (0.5) |
| Negative | 3,621 (99.3) | 134 (93.7) | 3,487 (99.5) | 31 (100) | 3,456 (99.5) | 25 (100) | 3,431 (99.5) |
| Contact density (median 3; IQR:1-4; Range 1-13) |  |  |  |  |  |  |  |
| ≤2 contacts *n(%)* | 769 (21.1) | 69 (48.3) | 700 (20.0) | 11 (35.5) | 689 (19.8) | 2 (8.0) | 687 (19.9) |
| 3 contacts *n(%)* | 849 (23.3) | 14 (9.8) | 835 (23.8) | 6 (19.4) | 829 (23.9) | 6 (24.0) | 823 (23.9) |
| 4 contacts *n(%)* | 808 (22.2) | 20 (14.0) | 788 (22.5) | 6 (19.4) | 782 (22.5) | 9 (36.0) | 773 (22.4) |
| ≥5 *n(%)* | 1,222 (33.5) | 40 (28.0) | 1,182 (33.7) | 8 (25.8) | 1,174 (33.8) | 8 (32.0) | 1,166 (33.8) |

*Abbreviations: HH – Household, IQR-Interquartile range, MTB-Mycobacteria Tuberculosis. \*percentages are all column percentages;*

### Contribution of repeat contact investigation to TB detection

Across all TB contact investigation time points, a total of 199 TB cases (79.9% bacteriologically confirmed) were detected, corresponding to an overall yield of 5.5% (n=199/3,648) (Table 3). Of these, 71.9% (n=143/199) were detected at baseline (Month 0), while 28.1% (n=56/199) were identified during follow-on screenings: 15.6% (n=31/199) at Month 6 and 12.6% (n=25/199) at Month 12. The TB detection yield declined progressively with each successive screening round, from 3.9% (n=143/3,648) at Month 0 to 0.9% (n=31/3505) at Month 6 and 0.7% (n=25/3,474) at Month 12.

**Table 2:** TB yields at different time points of the follow-on contact investigation cascade.

| Time point | Screened /investigated (N) | Total contacts diagnosed with TB, *n* (%)\* | Contacts diagnosed with bact. confirmed TB, *n* (%)\* |
|---|---|---|---|
| **Month 0** | 3,648 | 143 (3.9) | 106 (74.1) |
| **Month 6** | 3,505 | 31 (0.9) | 28 (90.3) |
| **Month 12** | 3,474 | 25 (0.7) | 25 (100) |
| **Total** | **-** | **199 (5.5)** \*\* | **159 (79.9)** |

\**percentages are all row percentages;* \*\**the denominator was total contacts evaluated at baseline*

## Discussion

This study identified a total of 199 TB cases (5.5%) among 3,648 household contacts of 1,250 bacteriologically confirmed index TB patients who were followed for one year. Most cases (71.9%) were detected at baseline, while 15.6% and 12.6% were identified at 6 and 12 months, respectively. These findings demonstrate the additional contribution of repeat contact investigation to TB case detection in a high-burden setting, despite the declining yield across successive rounds. Notably, nearly four in five (79.9%) of all detected cases were bacteriologically confirmed, which if detected late would contribute to ongoing household and community transmission. These results collectively highlight that intensive and longitudinal contact investigation can substantially increase case detection and help interrupt TB transmission in high-burden communities.

Our yield (5.5%) was notably higher than available evidence in similar low- and medium-income (LMICs) countries. Despite not accounting for the number of repeat TBCI rounds in each study, a recent systematic review and meta-analysis of contact investigation yield in LMICs, including 110 studies (about 39% from Africa), found a pooled active TB yield of approximately 2.87% (95% CI: 2.61-3.14) for all active TB (bacteriologically confirmed or clinically diagnosed), and about 2.04% (95% CI: 1.77-2.31) bacteriologically confirmed TB among contacts screened (14). Apart from one study conducted in Zambia that incorporated chest X-ray (CXR) with computer-aided detection (CAD) for screening household contacts and reported a 7.2% TB yield, albeit with a lower proportion of bacteriologically confirmed TB among all diagnosed TB cases (59.4%)(15), most studies from the region that employed more rigorous evaluation methods reported lower TB yields than observed in our study. In Uganda, a pilot project which optimized one time TBCI through strengthened home-visits, chest radiography and travel reimbursements only achieved a yield of 4.2%; majority (76%) of whom were clinically diagnosed based on X-ray findings (16). Moreover, a prospective cohort study in Ethiopia found no significant additionality between one time point TBCI in one site and a two-time repeat TBCI in another site: 1.1% vs 2.3% respectively (10). On the one hand, the smaller sample size (1, 276) relative to our study (3,648) may have limited the study’s statistical precision. Additionally, like other studies have focused on one-time screening or short follow-up, Wolde et al. conducted only two rounds of TBCI, which limited their ability to assess yield trends over time and capture additional TB cases that emerged after the initial screening (e.g., 12 months or more). Our inclusion of Months 6 and 12 is thus useful and could explain the higher overall yield. On the other hand, this could also reflect more intensive screening methods in our study at baseline and follow-on, high TB burden in the local population, or more comprehensive contact identification and follow-up.

In our study, TB detection yield declined sharply with each successive screening round (3.9% at Month 0 to 0.9% at Month 6 and 0.7% at Month 12). We observed that overall, there was a reliance on bacteriological confirmation at all screening time points: lowest at baseline (74%) and higher at follow on months: 90.3% at 6 months and 100% at 12 months likely missing clinical cases. This could partially elucidate the reduction in yield overall. Needless, the modest incremental yield in our setting aligns with findings from published meta-analyses showing that most active TB is detected early after exposure with most incident TB occurring within the first year post-exposure (14,17). Although TB detection yield declines with successive screening rounds, repeat contact investigations identify additional cases that might otherwise remain undetected, prolonging transmission and delaying treatment. Even modest residual incremental yields can have meaningful public health benefits in high-burden settings such as Zambia by enabling earlier diagnosis, treatment, and potentially improved outcomes(18,19). While the public health impact is evident, the economic justification for conducting repeat TBCI warrants further evaluation.

The sharp decline in yield over time suggests diminishing returns for general population contacts repeated follow-ups, raising the question of efficiency and cost-effectiveness. While baseline screening yields were high, lower yields at later rounds suggest the need for cost-effectiveness evaluation. Considering contact investigations and multiple follow-ups cost money and given the small incremental yield at later time points, policy makers need to weigh cost against benefit and decide the best implementation approaches.

Firstly, repeat contact investigations should be implemented strategically. For example, programs should consider targeting later screening rounds only to those at higher risk (e.g., contacts with co-morbidities, the elderly, young children) to optimize yield and cost-effectiveness. Additionally, disaggregation of yield by contact characteristics would inform targeting for follow-up TBCIs. Larger studies are urgently needed to identify contact characteristics that predict incident TB during subsequent rounds of repeat TBCI, to better inform targeted interventions. Furthermore, comprehensive cost-efficiency analyses should guide decisions on the optimal frequency of follow-up rounds, define priority contact groups, and determine whether integrating repeat TBCI into routine public health budgets is feasible or if sustained external investment is warranted. Secondly, because baseline screening relied primarily on symptom screening, some contacts with asymptomatic or minimally symptomatic TB may not have been identified during the initial investigation and only detected later during repeat TBCI after overt symptoms develop. More sensitive, rapid and accessible TB diagnostic tools should be used at month 0 to identify TB earlier, including asymptomatic or subclinical disease that might otherwise be detected only during subsequent screening rounds. This is supported by evidence from a Zambian household-contact study in which symptom screening alone would have missed 8 of 19 (42.1%) bacteriologically confirmed TB cases (15).

Combined with offering TPT to all eligible contacts in whom active TB has been excluded, more sensitive baseline screening may improve early detection, prevent incident TB among contacts with TB infection, and reduce reliance on resource- and labour-intensive, non-selective repeat TBCI. However, the effect of more sensitive baseline screening on the value of repeat investigation remains uncertain.

At the time of implementation, this intervention benefited from substantial operational support from the USAID-funded TB LON project, including the use of remunerated CBVs and project staff equipped with transport and airtime for follow-ups, even for contacts who had relocated.

This logistical capacity enabled complete follow-up and high retention of contacts in months 0, 6, and 12. However, such levels of resource availability may not be guaranteed under routine government-led implementation, where competing priorities and limited budgets often constrain active follow-up. Consequently, prioritizing contacts at highest risk of developing active TB after the initial investigation or enhancing baseline screening to identify asymptomatic or sub-clinical TB and TB infection, becomes essential for program sustainability and cost-effectiveness. These findings underscore the importance of integrating targeted screening and risk stratification int general health services to mitigate the challenges associated with resource constraints and ensure effective TB control.

This intervention had several limitations. The retrospective nature of the study and reliance on routinely collected programme data may have introduced documentation inaccuracies and biases. Although the two follow-up TBCI rounds identified additional TB cases, the study did not include the full recommended 24-month follow-up. Extending follow-up from 12 to 24 months after index-case detection could provide a more complete assessment of incident TB risk among household contacts. Another limitation was our inability to do chest x-rays on all contacts to screen for asymptomatic TB. Furthermore, we did not determine whether TB cases detected during repeat contact investigations were epidemiologically or genetically linked to their respective index cases. Phylogenetic studies are therefore needed to clarify transmission dynamics and establish whether TB cases identified during follow-up TBCI are linked to the presumed index cases.

## Conclusion

This study provides operational evidence that repeat tuberculosis contact investigation at 6 and 12 months identifies additional TB cases beyond baseline screening among household contacts in high-burden settings. Although detection yields declined across successive rounds, repeat investigations contributed 28.1% of all TB diagnoses. These findings highlight the potential value of longitudinal contact investigation while underscoring the importance of more sensitive baseline screening and risk-based follow-up strategies. Further research should evaluate the cost-effectiveness, optimal timing and targeting of repeat investigations to inform sustainable implementation in resource-limited settings.

## Data Availability

All data is submitted as part of this manuscript: aggregate, de-identified data underlying the analyses are publicly available as a supplementary dataset.

## Authors Contributions

Data extraction was done by Innocent Mwaba

Data collection and cleaning – Mutale Mukupa, Josaya Sakala and Innocent Mwaba

Data analysis – Daniel Siameka and Phallon Mwaba

The original draft of the manuscript by Phallon Blessing Mwaba

Critical review of the manuscript was done by Chitalu Chanda, Minyoi Mubita Maimbolwa, Muhammad Mputu, Nancy Kasese-Chanda, Kevin Mabvuto Zimba, Rhehab Chimzizi, Angel Mubanga, Mary Kagujje, and Monde Muyoyeta.

Supervision, validation and approval were done by Monde Muyoyeta and Mary Kagujje

All authors reviewed the manuscript and agreed to publish.

## Acknowledgements

The authors of this manuscript would like to acknowledge the staff at MoH health facilities who were involved in documenting patients and contact details in registers from where data was abstracted.

